# Do editorial board members publish in their own journal? – Review of editorial board members as authors in the main foot and ankle journals

**DOI:** 10.1101/2024.09.14.24312929

**Authors:** Manel Fa-Binefa, Albert Fontanellas Fes, Gemma González Lucena, Albert Ginés Cespedosa, Carlo Gamba

**Author notes:** Corresponding author: Gamba, Carlo; MD PhD Hospital del Mar (08003) Barcelona. Spain /93 248 33 35. **Level of evidence**: IV. **Declarations of interest**: None. **Funding**: This research did not receive any specific grant from funding agencies in the public, commercial, or not-for-profit sectors.

## Abstract

**Background:** Scientific publications by editorial board members in their own journals raise questions about potential biases in the peer-review process. This study investigates the prevalence of self-publishing in high-impact factor foot and ankle focused journals.

**Methods:** A review was performed of the highest impact factor foot and ankle focused journals/: *Foot and Ankle International*, *Foot and Ankle Surgery* y *Journal of Foot and Ankle Surgery.* In January 2023, the editorial board members were identified, and their names, roles, academic backgrounds, and regions were obtained. For each board member, the number of total published articles listed in PubMed and those for each corresponding journal over their entire history, during the last 5 years and during the last 3 years were compiled. Descriptive statistics analyses were performed using IBM SPSS Statistics v26.0.

**Results:** The study included 196 editorial board members from the 3 selected journals. Editorial board members have published 1694 articles in their own journals (9.17% of all the articles published in those 3 journals). Editorial board members published 23.4% (SD 23.6) of their overall production in their affiliated journals. Of that production, 39.0% (SD 38.6) have been published in the last 5 years, considering this 5-year period as the baseline for their role on the editorial board in question. Some 10% of editorial board members have published more than 50% of their scientific production in their own affiliated journal in the last 5 years. European editors (51%) have published more in their own journal over the last 5-year period than North Americans (29%) (p<0.001). Being a surgeon is related to greater publication rates in one’s own affiliated journal when compared to other specialist profiles (p=0.003).

**Conclusion:** The publication scientific articles by editorial board members as authors in journals with which they are affiliated is a present-day phenomenon in the highest impact factor journals with a focus on the foot & ankle. Knowledge of this data could be the key to understanding the prevalence of this phenomenon, and lead to making this data more accessible.

## 1. INTRODUCTION

The publication of articles in scientific journals is the main approach to communicating on research within the scientific community. Of course, it also serves as a means to gain recognition in scientific fields. Scientific publication is regulated by journal editorial boards, which oversee and control the quality of overall scientific literature available to all researchers in the scientific community. The ethical standards around this regulation play a key role in our seeing worthy scientific works that may influence future research and world decision making published. The import of those standards is greater in high-impact factor journals, with their broader coverage and influence. The tool available to editorial board to conduct this regulation is the Peer Review process. Despite the fact that the main scientific guidelines (ICMJE, COP, WAME) allow editors to publish in their own journals, there are recommendations to manage this complex occurrence and analyze its prevalence.[1]

Great variability in the self-publishing phenomena had been seen across different fields, journals and editors as referred to by many studies with distinct methodologies. [1] Medical journals have been reported to be almost three times more likely to publish reports from their own editorial board than is done in journals in other fields. [2] Moreover, the significant association between editorial board members and publication in their affiliated journals has been described in fields like General Surgery [3], oral health [4] and Urology [5]. To our knowledge, there is no published literature relative to the presence of this phenomenon in foot and ankle oriented journals.

The aim of this study was to analyze the frequency of the publication of articles by editorial board members of high-impact factor foot and ankle journals with which they are affiliated to understand its prevalence. Another aim was to increase our knowledge and understanding of this phenomenon to further continuing analysis of it. Our hypothesis is that there is data to support a finding of a significant association between editorial board members and publication in their own affiliated journals.

## 2. METHODS

### 2.1 Journal and author data

The research team contacted the editorial offices of all the journals via mail to obtain information on the structure of the editorial board, the changes in the composition of the editorial board as well as the date each editorial board member joined it.

The editorial board members of the main journals with highest impact factor with a focus on foot and ankle, according to Web of Science, available to publish scientific works without mandatory fee or invitation were included on the study. They are *Foot and Ankle International*, *Foot and Ankle Surgery* and *The Journal of Foot and Ankle Surgery*. *Foot and Ankle Clinics* was excluded as an invitation is required. *The Journal of Foot and Ankle Research* was excluded because of its mandatory publication fee. The editorial board members were identified from the official webpage of each journal in January 2023. Their full names, specialties (surgeon or non-surgeon) and regions (Europe, USA, Others) were obtained from the journal webpages. If the information was not available, searches on the main internet search engines were performed to obtain it.

### 2.2 Review

A review was performed during the first six-month period of 2023 by 2 researchers from 2 different institutions who were not on either of the editorial boards of the journals analyzed. A systematic search was performed in the PubMed database for all articles published by every editorial board member of *Foot and Ankle International*, *The Foot* and *Ankle Surgery* and *Journal of Foot and Ankle Surgery*. All comments on other publications or editorials were excluded from this search according to automatic search terms on the PubMed search engine (Example: NOT (Editorial [Publication Type]). For each case, searches were performed to obtain the total number of published articles in PubMed throughout its entire history (01-01-1900 to 31-12-2022), during the last 5 years (01-01-2018 to 31-12-2022), during the last 3 years (01-01-2020 to 31-12-2022) and during their period as an editorial board member if known. The time-period based searches were repeated to look for the number of articles published in the journal where they collaborated on the editorial board **(Figure 1).** In this way, the percentage of articles published in the journal for which they worked was obtained.

**Figure 1.**
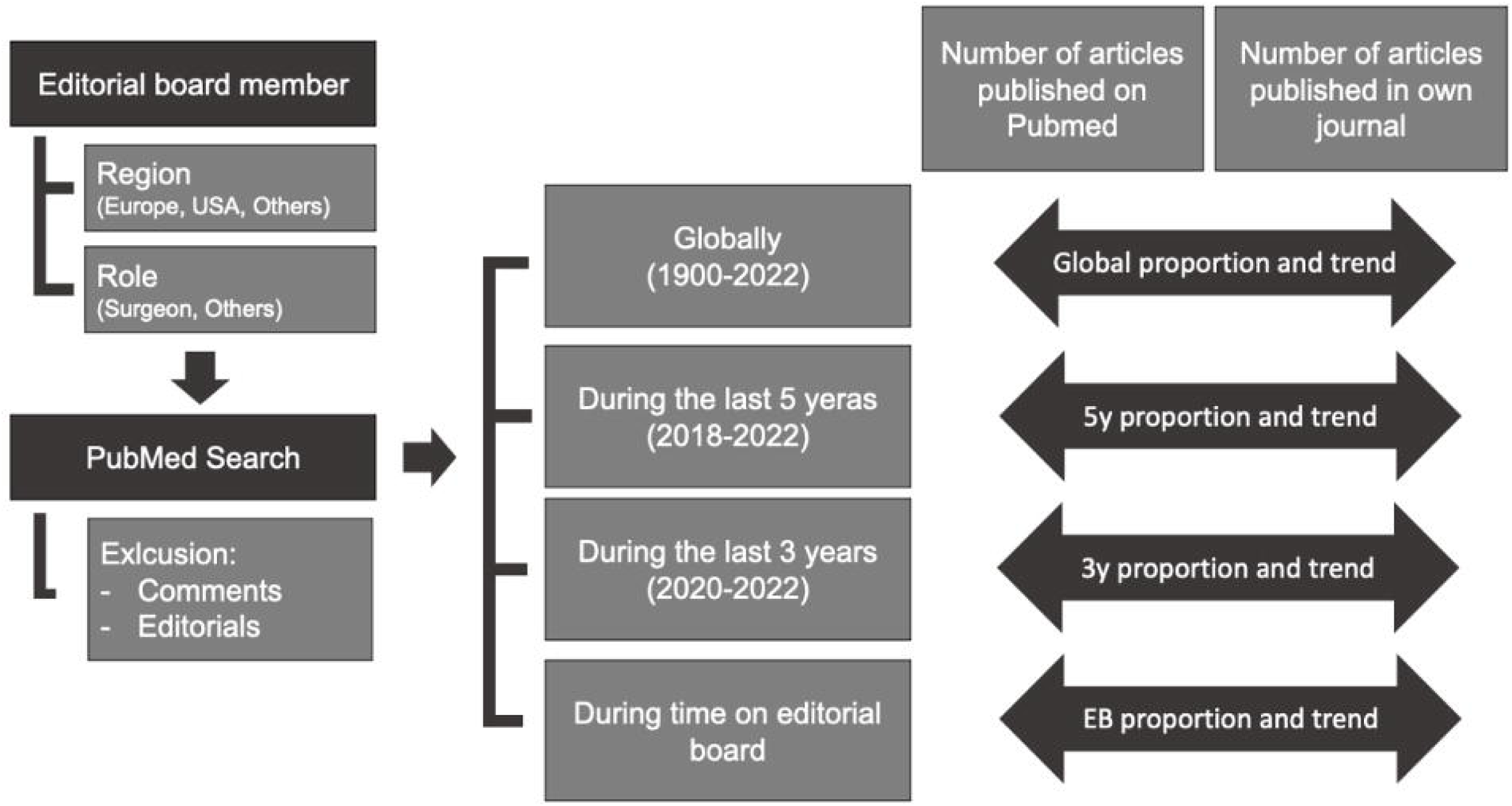
Flow chart of systematic search.

These timeframes (historical, 5-years and 3-years) were chosen by the authors in consideration of the a common/usual time for editorial board renewal and in case of failing to get information about the dates of editorial board member inclusion.

For those cases in which PubMed search results were confusing due to shared names with other authors, a special manual search was done by the researchers to identify the same data. It was first done with Orcid records if the information was available. It was not found in that source; it was done with the ResearchGate records.

Finally, for each journal, a new search was performed to obtain the number of published articles in PubMed during the same periods regardless of the author and excluding comments and editorials.

### 2.3 Statistical Analysis

The results were analyzed descriptively from the baseline database. Descriptive data was presented for all cases. For each case, the proportions between all published articles in the journals and the total number of articles published on PubMed during each period (history, last 5 years, last 3 years) were provided. Mean and standard deviations (SD) or median and interquartile ranges (IQR) were calculated for each period and journal. A trend ratio analysis was performed for each journal based on those results. Statistical analyses were performed with the IBM SPSS Statistics v26.0 (2019, Armonk, NY) software package.

## 3. RESULTS

### 3.1 Journal data

Editorial board make up is normally established based on the individual criteria of the editor-in-chief. The choice can be made from among reviewers based on quality of their work or it can be done *“ad personam”* based on their curriculum and knowledge. No information on the dates of adhesion to the editorial board of its constituent members was provided in any case and was not available on the official webpages of the journal. Editorial members renewal process, period and dates are not publicly available in any of the analyzed journals.

### 3.2 Baseline characteristics

The initial search provided 196 editorial board members from the 3 analyzed journals. The baseline characteristics of this group are shown in **Table 1**.

**Table 1.**
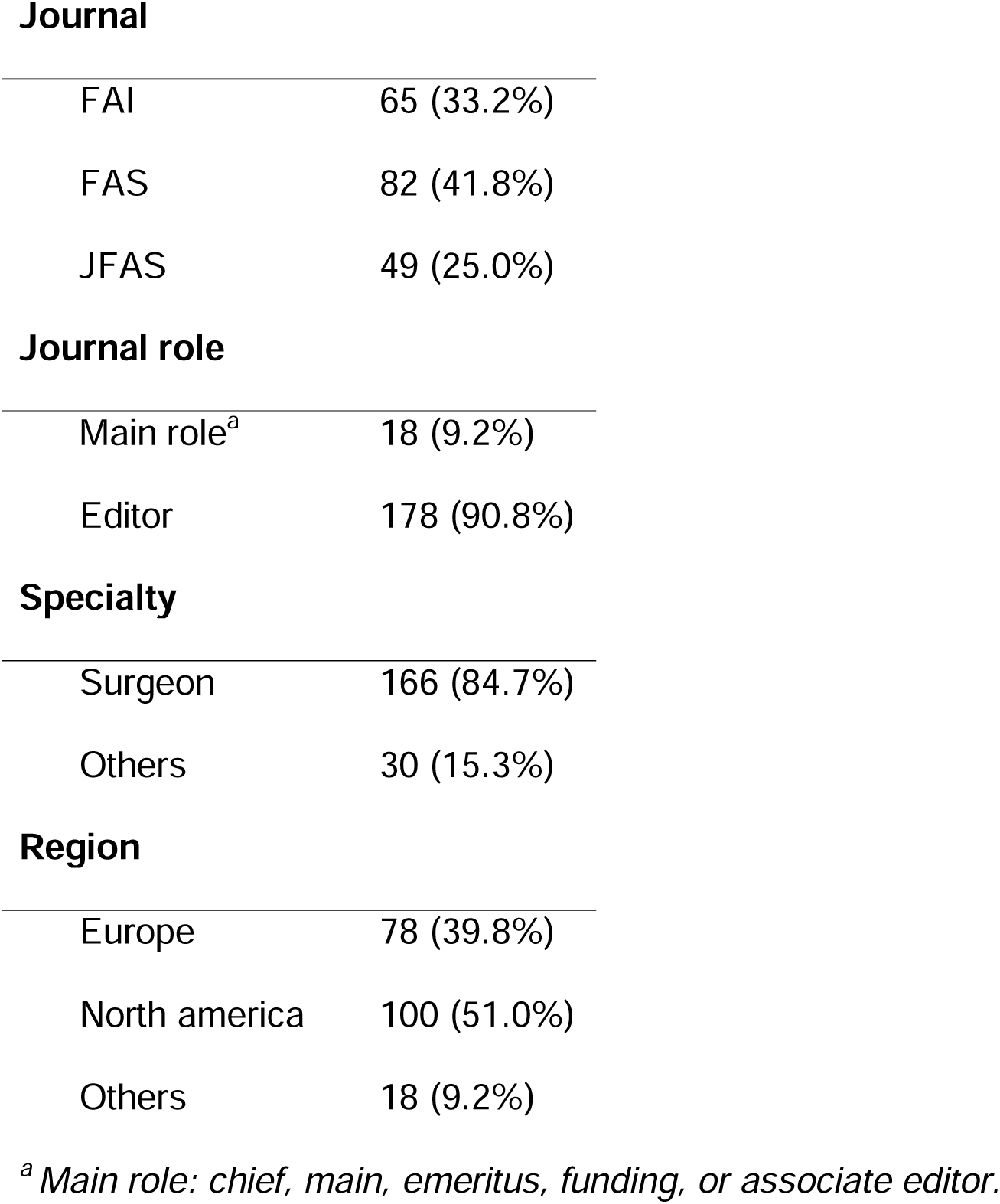
Baseline characteristics: number and % of editorial board members according journal, journal role, academic role, and region.

**Journal**
|  |  |
| --- | --- |
| FAI | 65 (33.2%) |
| FAS | 82 (41.8%) |
| JFAS | 49 (25.0%) |
| **Journal role** |  |
| Main role<sup>a</sup> | 18 (9.2%) |
| Editor | 178 (90.8%) |
| **Specialty** |  |
| Surgeon | 166 (84.7%) |
| Others | 30 (15.3%) |
| **Region** |  |
| Europe | 78 (39.8%) |
| North america | 100 (51.0%) |
| Others | 18 (9.2%) |

<sup>a</sup> Main role: chief, main, emeritus, funding, or associate editor.

### 3.3 Publication in own journal

The review of scientific publications by the 196 editorial board members on PubMed included a total of 1694 articles published in their affiliated journal throughout its history. They represented 9.17% of the total articles published in those journals. All the data of editorial members publications and its relationship with total PubMed publications and their own affiliated journal publications are presented in **Table 2**.

**Table 2.** Global data of actual editorial board members publications and its relationship with overall PubMed publications and own journal publications.

|  | FAI |  | FAS |  | JFAS |  |
| --- | --- | --- | --- | --- | --- | --- |
| Historically |  |  |  |  |  |  |
| Own journal articles (n) | 833 |  | 326 |  | 535 |  |
| In relation to all articles of same editors in PubMed (n, %) | 3438 | 0.24 | 3774 | 0.09 | 1491 | 0.36 |
| In relation to all articles of same journal (n/%) | 5358 | 0.16 | 8880 | 0.04 | 4230 | 0.13 |
| 2018-2022 |  |  |  |  |  |  |
| Own journal articles (n) | 286 |  | 211 |  | 163 |  |
| In relation to all articles of same editors in PubMed (n, %) | 1347 | 0.21 | 1587 | 0.13 | 371 | 0.44 |
| In relation to all articles of same journal (n/%) | 1038 | 0.28 | 3098 | 0.07 | 1297 | 0.13 |
| 2020-2022 |  |  |  |  |  |  |
| Own journal articles (n) | 169 |  | 136 |  | 98 |  |
| In relation to all articles of same editors in PubMed (n, %) | 820 | 0.21 | 1092 | 0.12 | 226 | 0.43 |
| In relation to all articles of same journal (n/%) | 593 | 0.28 | 2089 | 0.07 | 814 | 0.12 |

Editorial board members have published 23.4% (SD 23.6) of their global production in their own journal and of this production, 39.0% (SD 38.6) has been published in the last 5 years, considering this time lapse related to their role on editorial board. Editorial board members publications in their own journals in comparison to all PubMed published articles increased from 17.66% in the time frame from 1899 to 2018 to 22.14% in the time frame from 2018 to 2022. Ten percent of the editorial board members have published more than 50% of their scientific production in their own journal in the last 5 years. Up to 13% of all-history articles published in a journal include actual editorial board members among the authors. The descriptive analysis of scientific publications according to region and journal are shown in **Table 3 and 4**. European editors have published more in their own journals in the last 5 years (51%) than North Americans (29%) (p<0.001). Surgeons have been observed to publish more articles in journals where they hold editorial roles when compared to other specialist roles. This trend was statistically significant over the past 5 years (p = 0.003) and 3 years (p = 0.008). Furthermore, when compared to the total number of articles published on PubMed in the last 3 years (p = 0.01), 5 years (p = 0.001), and historically (p < 0.001), the publication rates of surgeons in their own journals remained significantly higher. In terms of journal roles, holding a primary position (such as chief, main, emeritus, funding, or associate editor) did not show a statistically significant difference in the rate of publishing in one’s own journal when compared to other editorial positions (p = 0.178).

**Table 3.**
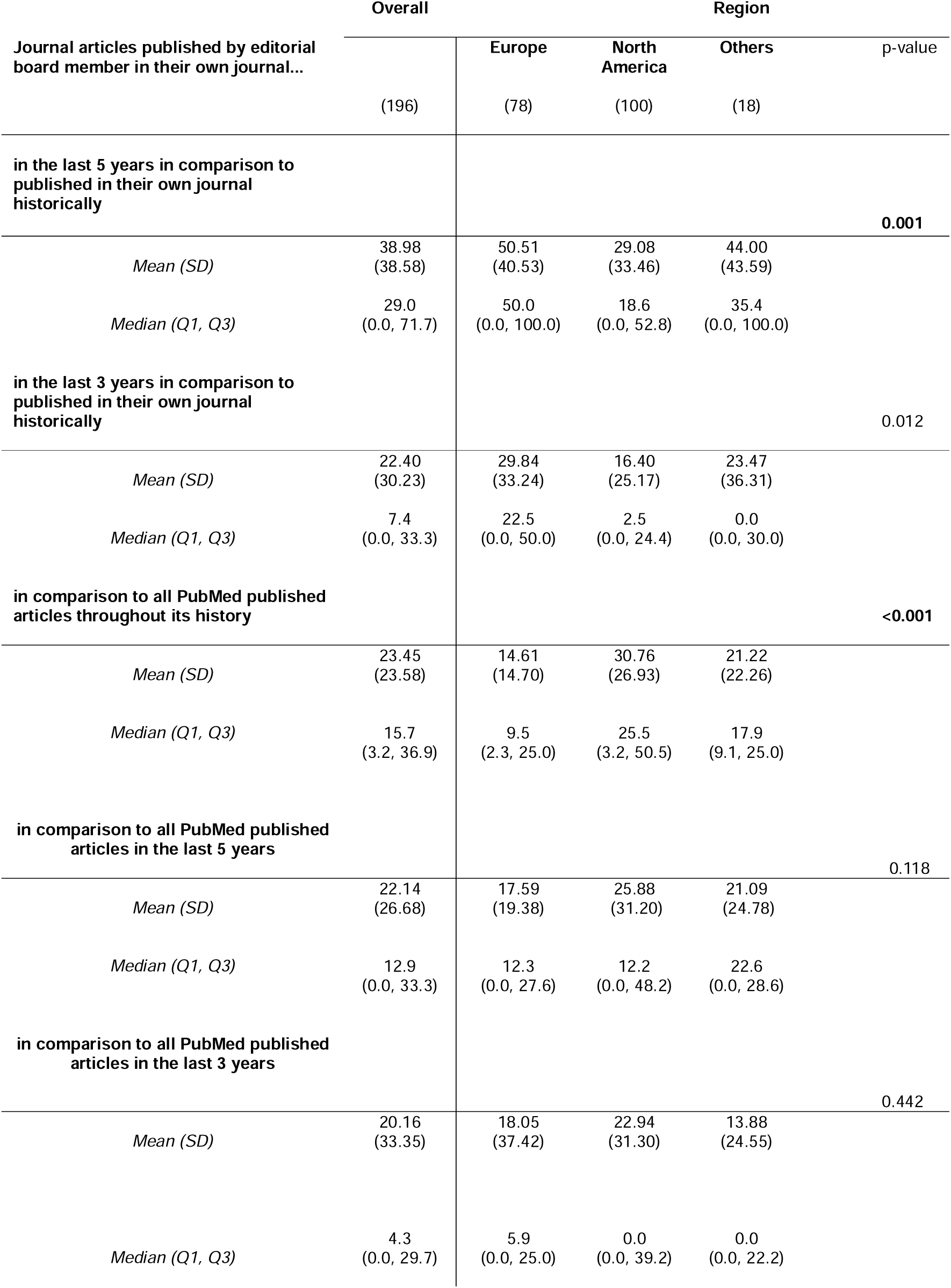
Editorial board members publications according to the region of the editorial board members.

**Table 4.** Editorial board members publications according to journal of editorial board members.

| Journal articles published by editorial board member in their own journal... | Overall | Region |  |  | *p*-value |
| --- | --- | --- | --- | --- | --- |
|  |  | Europe | North America | Others |  |
|  | (196) | (78) | (100) | (18) |  |
| **in the last 5 years in comparison to published in their own journal historically** |  |  |  |  | **<0.001** |
| *Mean (SD)* | 37.48<br>(33.84) | 37.48<br>(33.84) | 50.11<br>(43.16) | 22.32<br>(29.65) |  |
| *Median (Q1, Q3)* | 28.8<br>(0.0, 62.5) | 28.8<br>(0.0, 62.5) | 50.0<br>(0.0, 100.0) | 0.0<br>(0.0, 40.0) |  |
| **in the last 3 years in comparison to published in their own journal historically** |  |  |  |  | **0.017** |
| *Mean (SD)* | 20.23<br>(25.84) | 20.23<br>(25.84) | 29.10<br>(35.83) | 14.06<br>(22.37) |  |
| *Median (Q1, Q3)* | 12.5<br>(0.0, 27.9) | 12.5<br>(0.0, 27.9) | 0.0<br>(0.0, 50.0) | 0.0<br>(0.0, 20.0) |  |
| **in comparison to all PubMed published articles throughout its history** |  |  |  |  | **<0.001** |
| *Mean (SD)* | 26.15<br>(21.41) | 26.15<br>(21.41) | 14.06<br>(16.98) | 35.61<br>(29.13) |  |
| *Median (Q1, Q3)* | 23.8<br>(10.0, 36.4) | 23.8<br>(10.0, 36.4) | 9.1<br>(1.1, 21.4) | 42.9<br>(1.1, 53.3) |  |
| **in comparison to all PubMed published articles in the last 5 years** |  |  |  |  | **0.039** |
| *Mean (SD)* | 22.76<br>(21.71) | 22.76<br>(21.71) | 17.28<br>(21.51) | 29.47<br>(37.23) |  |
| *Median (Q1, Q3)* | 20.0<br>(0.0, 33.3) | 20.0<br>(0.0, 33.3) | 8.7<br>(0.0, 25.8) | 0.0<br>(0.0, 60.0) |  |
| **in comparison to all PubMed published articles in the last 3 years** |  |  |  |  | **0.109** |
| *Mean (SD)* | 18.00<br>(20.91) | 18.00<br>(20.91) | 16.71<br>(37.63) | 28.80<br>(38.00) |  |
| *Median (Q1, Q3)* | 11.1<br>(0.0, 30.0) | 11.1<br>(0.0, 30.0) | 2.6<br>(0.0, 25.0) | 0.0<br>(0.0, 66.7) |  |

## 4. DISCUSSION

Editorial board members of high-impact factor foot and ankle focused journals publish 9-36% of their scientific publications in their own journals. That figure has increased to 21-43% in the most recent 5 years, relating this period to their being-in-charge period on the editorial board. These publications represent between 4-28% of the total articles of those journals. This work also saw significative differences in publication trends depending on the specialty of the editor, region of the editor, and the journal. Surgeons generally publish more in the journal with which they are affiliated than other specialty profiles publishing in foot and ankle journals. European editors have published more in their own journal in the last 5 years in comparison to previous years (50.51%, p=0.001) while North American editors published more in their own journal than in other journals historically (30.75%, p<0.001). This data is in line with previous research in other fields showing the “Self-publishing” phenomena at up to 7% in other fields[6], 1-25% in medical specialty journals [2] and 2-27% in surgical peer-reviewed publications. [3]

The limitations of this review include the use of a single scientific database, the lack of authorship authentication systems and the lack of data on the year of adhesion to the editorial board. In this study a single scientific database (PubMed) has been used, which can bring a bias to the results since some publications are only available in other databases. However, due to amount of analyzed data and quality of analyzed journals, publications not available on PubMed would probably be scarce and its effect on the results presented should be minimum. Another limitation is the lack of available authorship authentication systems and the use of different names by some authors. Moreover, an overestimation may be present in cases in which two or more members of the editorial board publish together as the same article may appear duplicated in our search. This phenomenon could even bring on a more difficult and complex ethical debate and must also be considered. Finally, the lack of available homogenous data on the editorial board adhesion year of its members is also an important limitation. That one was overcome with the assumption of a recent 5-year period as related to an editorial member position. The strengths of this review include being the first analysis of its characteristics in Orthopedics, and in the foot and ankle field. Despite this research being a survey of some of the main foot and ankle journals, this phenomenon is probably also present, or even more present in journals, with a lower impact factor with maybe less availability of ethics and authorship control mechanisms. The main ethics guidelines on scientific publication suggest applying mechanisms to control editors publishing in their own journals.[7,8] Regarding the results of this work, some mechanisms that can increase journal transparency and the availability of this data and its analysis could be journals providing clear available information on editorial board member adhesion dates and the years in charge of every editorial board member. They would be a simple option to increase transparency on the issue. Another option would be for journals to make an authorship authentication system mandatory for editorial board members, such as ORCID.

## 5. CONCLUSION

The publication of scientific articles of editorial board members as authors in their own journals is a present-time phenomenon in the highest impact factor foot and ankle journals and it represents an important component of the global scientific production of these authors. A greater knowledge and awareness of this data and phenomena could be key to understanding its prevalence, handling these data with transparency, and, if necessary, performing further analysis and/or specific control.

## Data Availability

All data produced in the present study are available upon reasonable request to the authors.

## REFERENCES

[1] Helgesson G, Radun I, Radun J, Nilsonne G. Editors publishing in their own journals: A systematic review of prevalence and a discussion of normative aspects. Learn Publ 2022;35:229–40. 10.1002/leap.1449.

[2] Luty J, Arokiadass SMR, Easow JM, Anapreddy JR. Preferential publication of editorial board members in medical specialty journals. J Med Ethics 2009;35:200–2. 10.1136/jme.2008.026740.

[3] Sen-Crowe B, Sutherland M, Shir A, Kinslow K, Boneva D, McKenney M, et al. Variations in surgical peer-reviewed publications among editorial board members, associate editors and their respective journal: Towards maintaining academic integrity. Ann Med Surg 2012 2020;60:140–5. 10.1016/j.amsu.2020.10.042.

[4] Rösing CK, Junges R, Haas AN. Publication rates of editorial board members in oral health journals. Braz Oral Res 2014;28:1–5. 10.1590/1807-3107BOR-2014.vol28.0042.

[5] Mani J, Makarević J, Juengel E, Ackermann H, Nelson K, Bartsch G, et al. I publish in I edit?--Do editorial board members of urologic journals preferentially publish their own scientific work? PloS One 2013;8:e83709. 10.1371/journal.pone.0083709.

[6] Zdeněk R. Editorial Board Self-Publishing Rates in Czech Economic Journals. Sci Eng Ethics 2018;24:669–82. 10.1007/s11948-017-9922-2.

[7] Committee On Publication Ethics (COPE). Best practice guideline for journal editors. 2022.

[8] Committee On Publication Ethics (COPE). A short guide to ethical editing for new editors 2022.

